# FATAL: A Forensic AuTopsy Annotation tooL for digital recording of autopsy findings

**DOI:** 10.1101/2024.02.23.24303284

**Authors:** Mikkel V. Petersen, Asser H. Thomsen, Kasper Hansen

## Abstract

The findings from forensic autopsies, where cause of death must be established and reported to legal authorities, are reported in paper-based formats. Practitioners are required to map 3D injury findings to 2D space. Here, we design and describe a digital Forensic AuTopsy Annotation tooL (FATAL), that can be used by practitioners to record systematically detailed autopsy findings onto an interactive 3D body model.

We employ a user-centred design process involving an expert forensic medicine team. We describe the iteration process and the final functionality determined, based on in-depth analyses of forensic clinical workflows, and feedback on the types of complex cases confronting practitioners.

FATAL functions include freehand drawing, a layer system for injury categorisation, trajectory plotting, surface area markings, and point-of-interest marking. Relevant external images, such as investigative report or autopsy photographs, can be loaded into the FATAL tool and assigned to individual annotations. The application streamlines workflows, supports template-driven documentation, and collates all forensic data into a single interface. Findings from the digital tool can be exported to a 2D report (PDF).

We highlight the advancements in accuracy, efficiency, and reproducibility afforded by a digital tool for forensic autopsy documentation. Potential applications in forensic medical examinations beyond autopsies are described, along with specific areas for extension, such as supporting touch screen and pen inputs, export for 3D printing models and extending the tool’s compatibility with custom 3D body models.

**Highlights:**

- FATAL is a digital platform for systematised detailed documentation of autopsy findings
- The platform enables accurate annotation of injuries onto a 3D body model
- Allows the forensic pathologist to move from traditional paper-based body diagrams to interactive 3D visualisations
- The platform can export annotations to 2D body diagrams and embed them directly in custom PDF templates

## 1. Introduction

Forensic autopsies are mandated by law in many countries, where the cause of death is suspected to be non-natural [1] and provide requisite evidential data for medico-legal cases. In contrast to the falling numbers of clinical autopsies worldwide [2, 3], forensic autopsy rates remain stable [4]. Despite their mandatory status and consistent numbers year on year, there has been relatively little change to forensic autopsy procedures over the past decades. Typically, a forensic autopsy requires that a forensic pathologist or specialist (MD) in forensic medicine documents the exam findings onto a paper-based two-dimensional (2D) model of a body identifying injuries and deformations.

Documentation of a three-dimensional body to a two-dimensional paper-based model has inherent challenges [5]. Accurate documentation requires extensive training, anatomical knowledge, and precise notations can vary from pathologist to pathologist [6]. Digital technologies offer clear opportunities to streamline and systematise procedures beyond what is possible with paper-based methods and might ultimately improve the accuracy and precision of documentation [7, 8, 9]. For example, rendering the results of an autopsy examination with a digital platform could allow a pathologist to interact with a three-dimensional model of the body, zoom in and out of important anatomical regions and use standardised annotation tools. Amendments to annotations can also be made more convenient versus paper-based formats, along with exporting finalised body models to either a traditional PDF or a three-dimensional model for further rendering.

There has been recent interest in innovations to forensic procedures afforded by immersive imaging technologies [10]. Much of this work has focused on augmenting existing physical formats with data rendering or developing virtual autopsy formats. For example, one study demonstrated the use of augmented reality headsets to render post-mortem computed tomography (CT) images during autopsy examinations [11, 12]. Other work developed a prototype of a mixed-reality system to present whole-body visualisations, CT images and other data such as police reports, with the ultimate goal of replacing physical autopsy [5].

Despite the developments in immersive technologies, there has been relatively little work on digital tools for autopsy documentation. Improving the efficiency, cross-team collaborative capacities and even training within forensic autopsy workflows are important, given the shortage of forensic scientists in many countries [13]. Therefore, in this work, we design and describe a digital Forensic AuTopsy Annotation tooL (FATAL), that can be used by pathologists to record systematically detailed information on autopsy findings onto a 3D body model. The goal of this work is to describe the requirements identified for a digital tool, from the initial opening display to data export, and the useful functionality determined over multiple rounds of iteration with expert forensic scientist users.

## 2. Method

### 2.1. Overview of FATAL

FATAL is developed as a standalone desktop application for Windows 10/11. We used the cross-platform game engine Unity (Version 2023.2.1f1) for application development.

We developed the application iteratively, with a user-centred process beginning with interviews and proceeding through 7 rounds of software iterations and 4 in-person meetings discussing application features. Throughout the development process, two researchers at the Department of Forensic Medicine (MD/PhD specialist in forensic medicine; MSc/PhD forensic researcher) provided formative inputs to the FATAL application, which was developed by a researcher at the Department of Clinical Medicine (MD/PhD). In Denmark, the medical system trains physicians to be “specialist in forensic medicine”, which broadly corresponds to international designations of “forensic pathologists”. To be consistent with the international studies, we use the term “forensic pathologist(s)”, the intended user group of the annotation tool”

### 2.2. Development Model

The first stage of development comprised an initial interview, where the specialist in forensic medicine described local procedures and workflows for autopsies. The hour-long interview focused on current paper-and-pencil methods for autopsy annotation. In particular, we discussed the challenges faced by forensic pathologists in drawing and communicating the trajectories of injuries (e.g., stab wounds) using current non-digitised formats. Figure 1 presents an example case with the paper forms used by the forensic medicine team in the local hospital setting. These forms were formative in discussions of the core functionality required.

**Figure 1:**
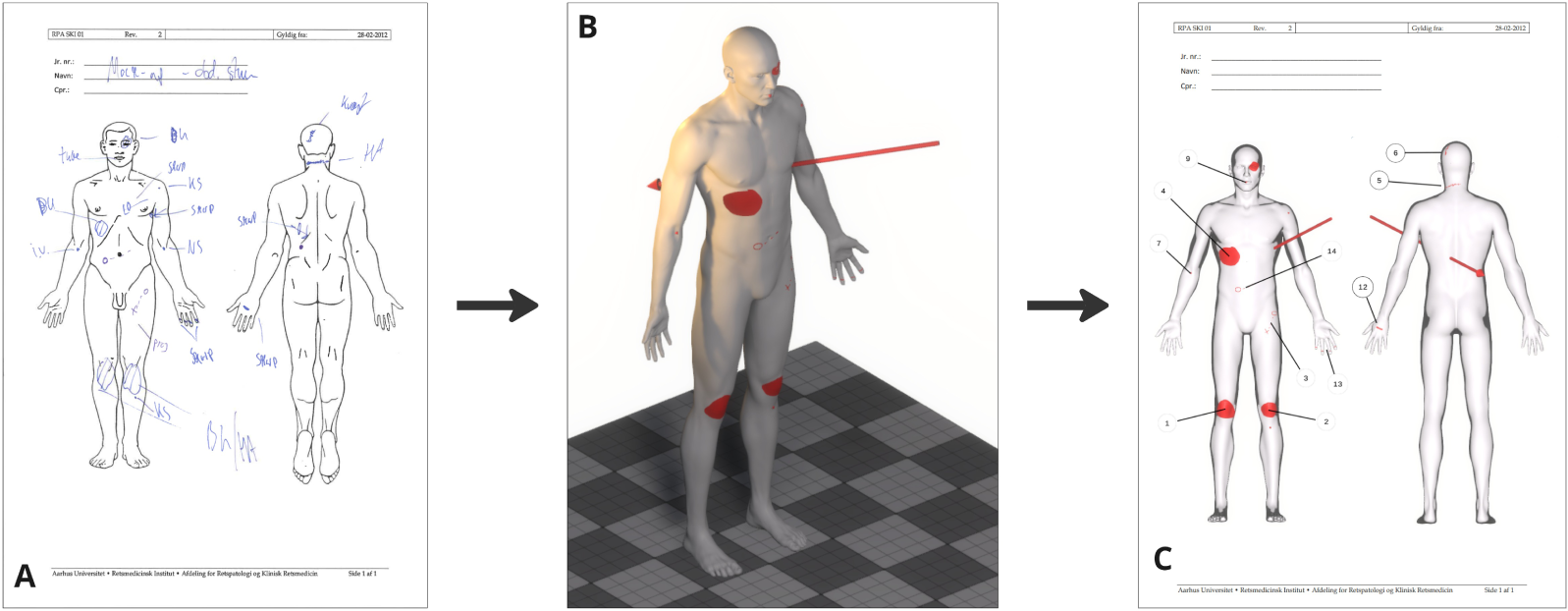
A) Mock-up forensic case (not actual patient data) with hand-drawn annotations on paper form used at the local department. B) 3D view of annotations registered in the FATAL app. C) Example of a 2D PDF report exported directly from the FATAL app.

From the first interview, a basic prototype was developed, illustrating how wound trajectories might be plotted digitally, with the aim of exporting to a model for 3D printing. The initial prototype stimulated further discussions on appropriate data presentation (e.g., the type of body model presented, level of detail and rendering), and discussions on the mathematical calculation of wound angles, and the data outputs on angles. We then scheduled a series of physical interviews to explore further possibilities emerging from initial prototype feedback. After each physical interview (five in total from September 2022 until November 2023), the prototype was further iterated and updated. Additional feedback on functionality, including corrections to function implementations, perceived usefulness and further development was provided via email. The specialist in forensic medicine also developed several paper mock-ups of a sample autopsy (Figure 1A) to illustrate the challenges he typically faces, and to further clarify how he might approach specific problems.

In total, 7 prototype versions of the application were deployed to the two expert users. To enable high-frequency updates, a custom app launcher (Windows 10/11) was built using Windows Presentation Foundation and a backend Web API (Python, FastAPI) was deployed to automatically push updated application builds to the end users. The aim was to shorten the developer-user feedback loop.

### 2.3. Platform Requirements

Table 1 presents key outputs from the iterative design process, along with the end-stage list of requirements for functionality. A number of the iterations focused on bringing the digital tool closer to the paper-based annotation experiences of the forensic pathologist. Flexibility of annotation emerged as a key requirement (e.g., replicating the experience of free-hand drawing) because of the diversity in injuries encountered in forensic medicine, and body points of interest (e.g., bruises, scars) that might be noted during the autopsy. In the final iteration, more complex requirements, such as layer grouping, were implemented, allowing more sophisticated rendering than possible in paper formats.

**Table 1:**
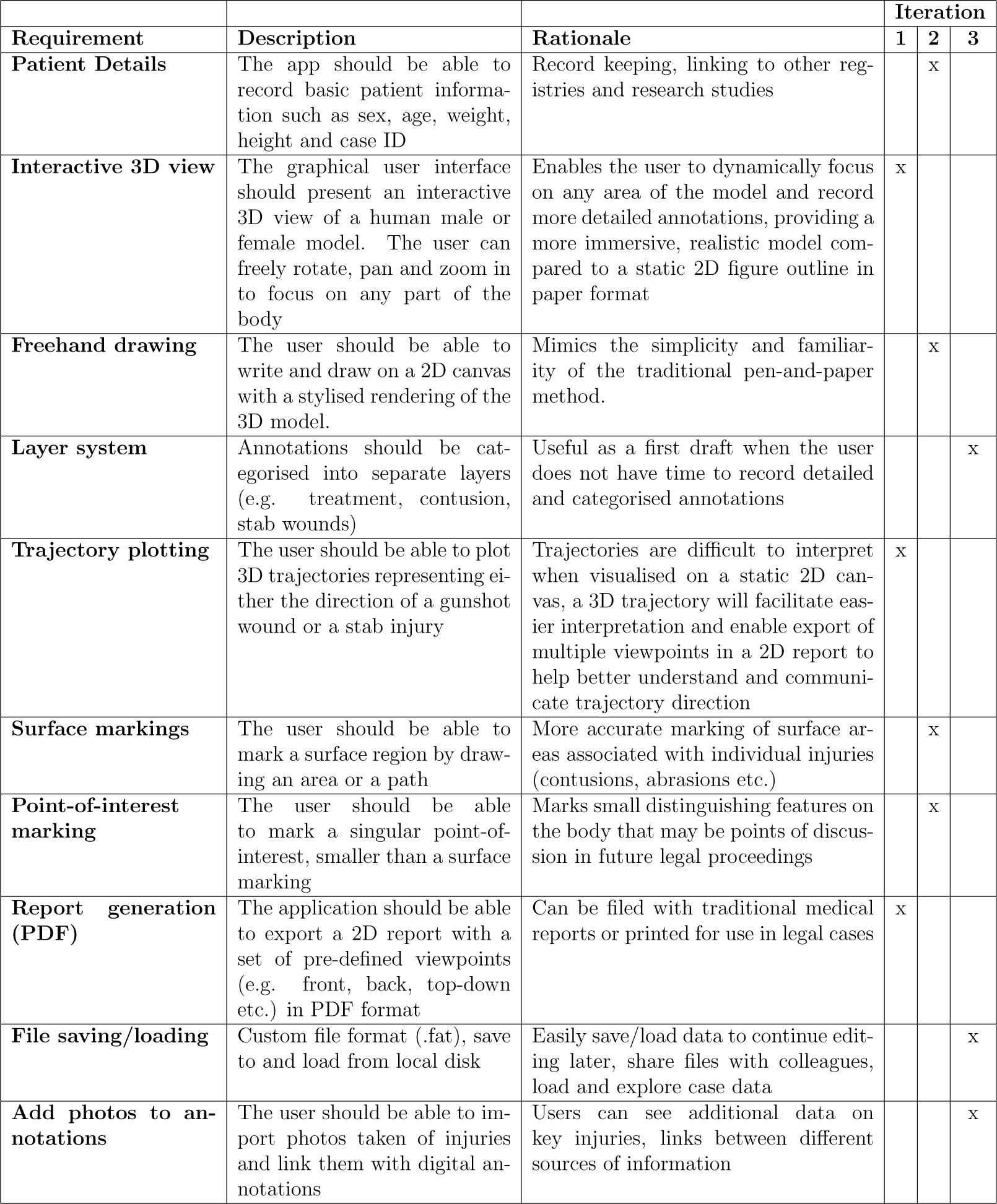
Table listing platform requirements, feature descriptions and the rationale for their implementation.

## 3. Results

The core features of FATAL are outlined in Figure Figure 2.

**Figure 2:**
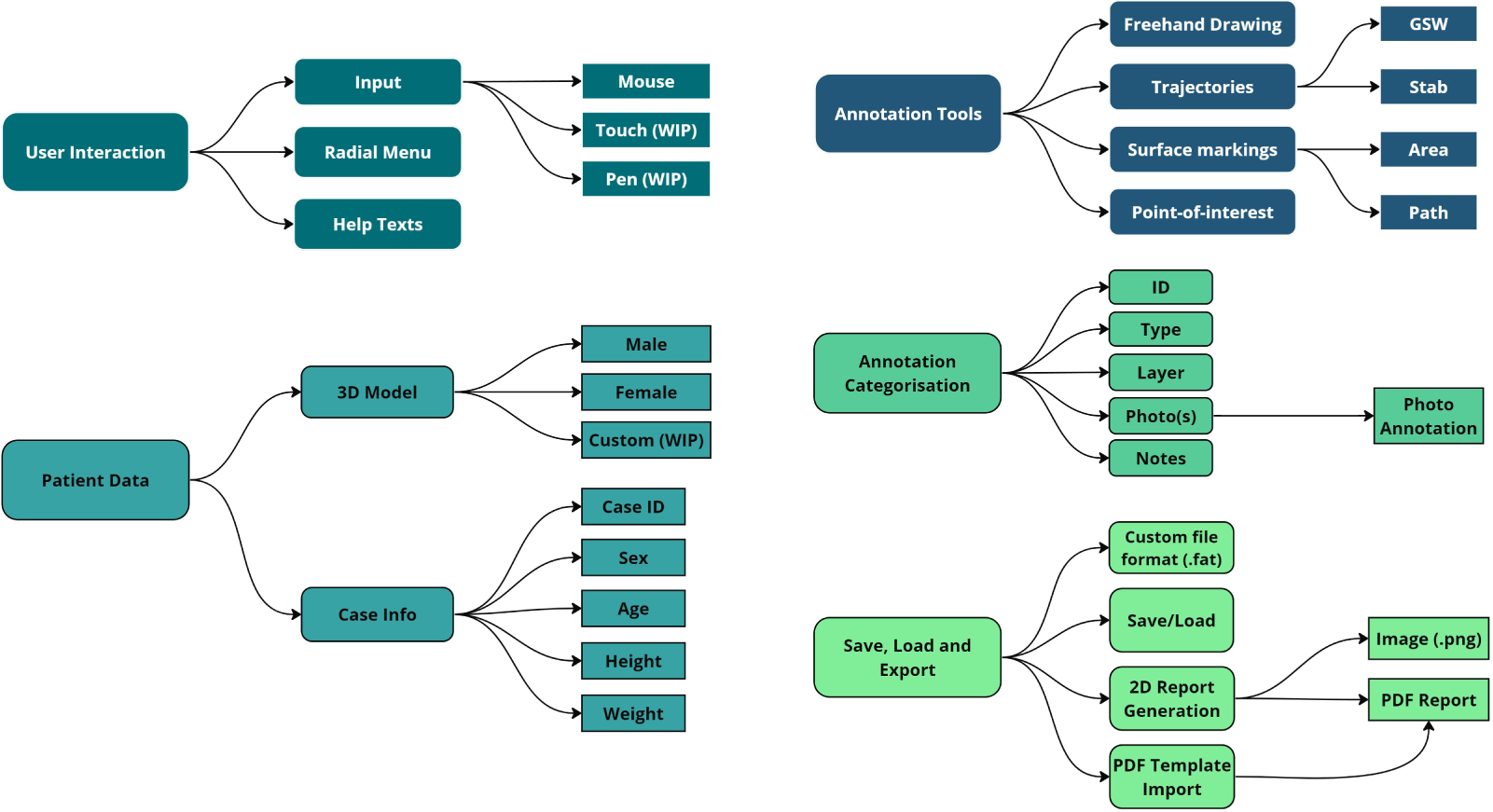
Overview of the FATAL app’s core features and implemented data types. (WIP) Work-In-Progress.

### 3.1 Workflow

The primary aim is to ensure a simple user experience with intuitive menus to help forensic pathologists get started with the tool. First, patient details are inputted by the user, and then an interactive three-dimensional (3D) body model is presented.

As a default, the application presents text instructions to guide users, particularly those new to the tool. The instructions can be disabled in the settings menu when the user feels they are no longer useful. The application can be set to dark mode to reduce eye strain if used in the later hours.

### 3.2 User Interface

Figure 3 provides an overview of the graphical user interface across different modes.

**Figure 3:**
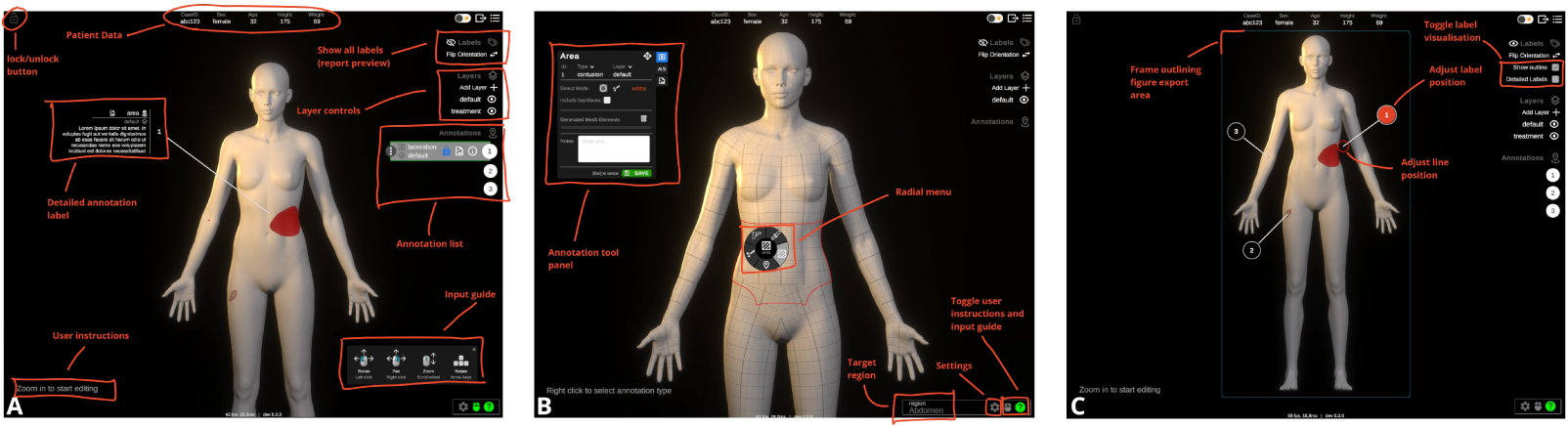
Overview of the application user-interface (UI) in different modes. A) View Mode, B) Edit Mode, C) Report Preview. Key UI functionality outlined in panels.

The application in “view mode” allows the user to pan, zoom and rotate the body model in three-dimensional space. In addition to the 3D body model, this view will also visualise annotations placed on the surface of the model. Individual annotations can be selected, either directly on the model or via the list of annotations in the user interface (UI), such that it is highlighted with its reference label (default is the annotation ID) and additional annotation details displayed (e.g. type, layer, notes, photos). All annotation labels can be visualised simultaneously in the ‘report preview’. Here the visualised labels can be filtered by individual layers (e.g., contusions, gunshot wounds (GSW), treatment).

The report preview visualises the canvas area that is exported (rectangular outline) and how the labels will be positioned in the generated PDF report. The preview is locked to either a front- or back-facing view. Individual annotation labels can be manually re-positioned on the 2D canvas to ensure optimal positioning (i.e., so that label lines are not covering other labels or annotations).

If a previously annotated case is loaded (custom .fat file), the app will start in view-only mode and will need to be unlocked before editing or adding annotations. When unlocked, “Edit mode” is enabled automatically when the user zooms in on an anatomical region. A mouse right-click opens a radial menu (Figure 3B), allowing the user to select one of five annotation modes (freehand drawing, GSW-trajectory, stab-trajectory, Area and Point-of-interest). After selecting an annotation mode, a mode-specific UI panel opens (Figure 3B).

### 3.3 Patient data

A male or female body model can be selected. The 3D models provide a basic template of male and female anatomy, derived from models created by Andrei Cristea (2022) (https://lhndo.github.io/undoz/blog.html, retrieved on: February 20th, 2024). The original models were manually processed in Blender (Blender Foundation, https://www.blender.org/) to place them in the standard anatomical position for annotation and to subdivide them into anatomical regions.

Patient demographic details including sex, age, height and weight can be entered in addition to an auto-generated case ID. The generated case ID can be edited to match any internal departmental system for patient identification.

### 3.4 Annotation Tools

An overview of the implemented annotation tools is provided in Figure 4.

**Figure 4:**
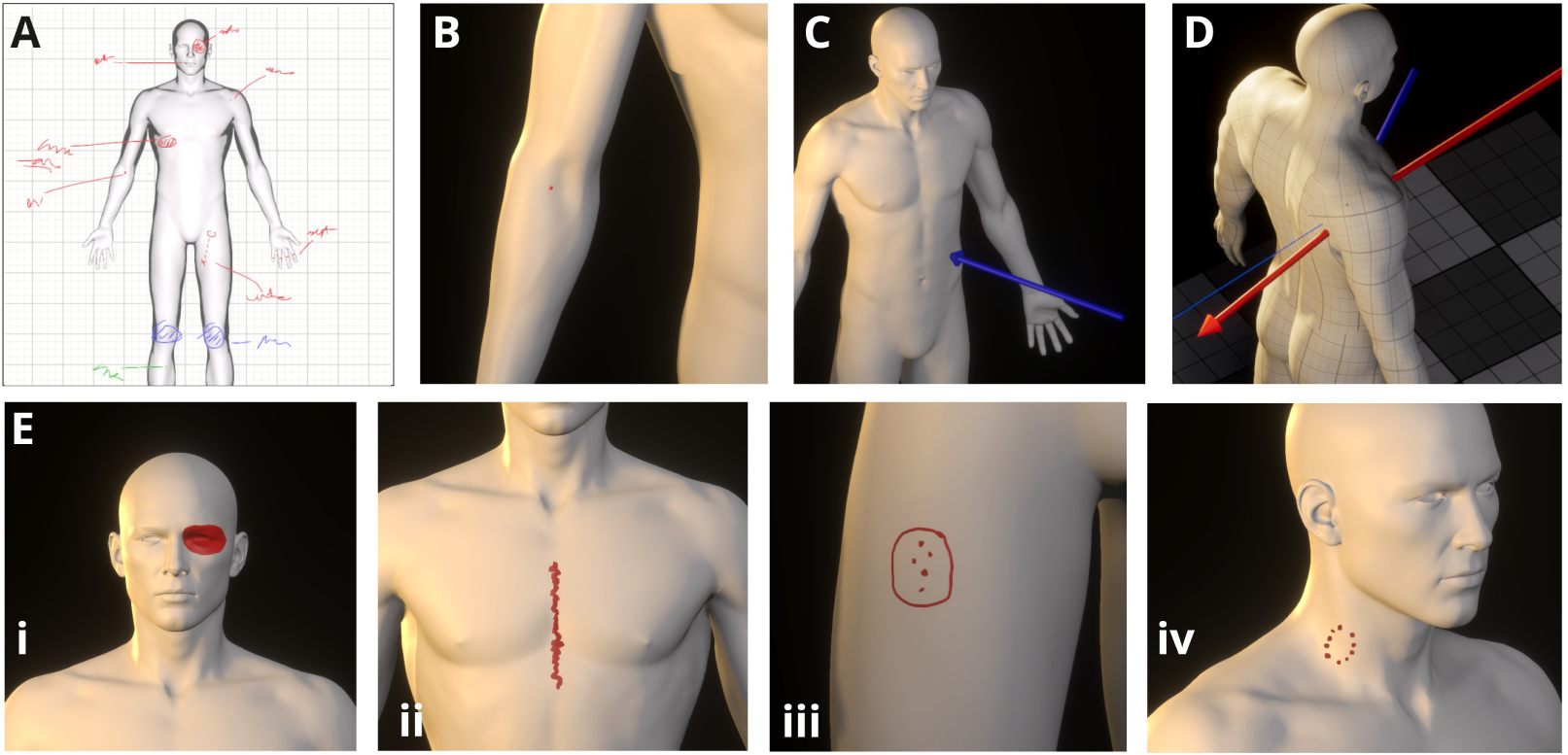
Overview of the different annotation tools. A) Freehand drawing, B) Point-of-interest (e.g. needle marks), C) Stab-trajectory (non-perforating), D) GSW-trajectory (perforating with deflected exit trajectory), E) Surface markings (i) Closed-loop area, (ii) Open-loop path, (iii-iv) multiple surface markings grouped in a single annotation.

#### 3.4.1 Freehand drawing

A freehand drawing module is implemented so that forensic pathologists retain the simplicity and familiarity of traditional paper-based annotations (Figure 5). Using the digital freehand drawing provides the user with additional flexibility allowing them to change pen colour and thickness and allows for easy editing with undo, redo and erasing functionality.

**Figure 5:**
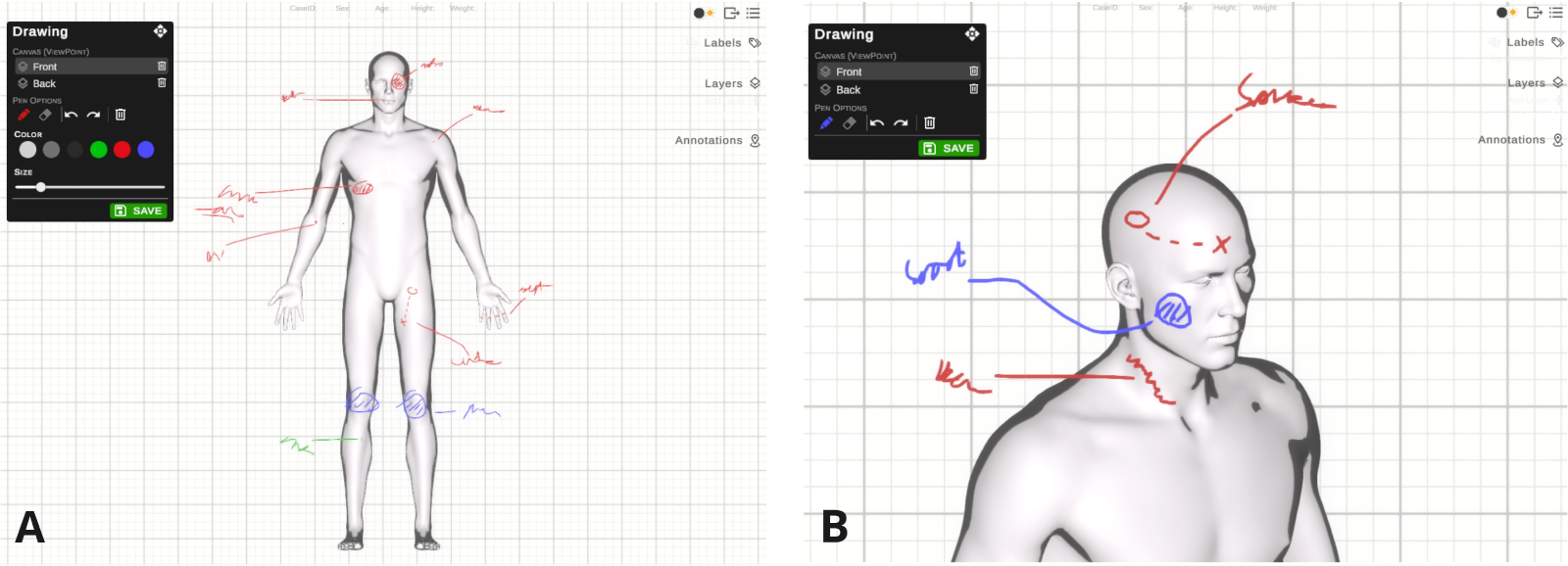
Freehand drawing. A) Overview of the freehand drawing UI, B) Example of oblique view allowing more accurate annotation drawing.

A major constraint of paper formats is the limited viewpoints available: annotations can be drawn on a front or rear-facing body. The present tool addresses this constraint by allowing the user to select any possible viewpoint, dynamically rendering a figure outline, to facilitate accurate drawing of injury outlines and notes.

The freehand drawing model module can be used to make quick notes during the autopsy. These sketch notes can be used as a first draft before later converting them to categorised annotations, using other functionality of the application.

#### 3.4.2 Surface area marking

The surface marking module is implemented to allow the user to annotate surface areas on the 3D model for specific injuries (Figure 4E). The module has two modes: area and path. In “area mode” the user draws a closed-loop circle, and the application will extract and colour the area. In “path mode”, the user can select a line width and draw a path (open-loop) on the surface of the model (as opposed to the freehand drawing mode, where the paths are placed on a 2D canvas with a projected figure outline).

Multiple surface areas and paths can be grouped in a single annotation, allowing the user to draw more complex patterns (e.g., bite marks or close range and contact gunshot wounds) or to group related injuries (e.g., needle marks) (Figure 4E).

#### 3.4.3 Trajectories

Trajectories are subdivided into (1) gunshot wounds (GSW) and (2) penetrating stab wounds (Figure 4C+D). Both trajectory types can be either perforating or non-perforating. If a GSW is perforating, then the exit trajectory can be controlled separately from the entry trajectory, in case of deflection of the bullet when passing through the body. If the pathologist can determine the depth of a non-perforating GSW or stab wound, this can also be recorded in the trajectory annotation.

Trajectories are defined in FATAL using (1) a forward or backward direction, (2) a horizontal angle (0-90 degrees towards the left or the right) and (3) a vertical angle (0-90 degrees downwards or upwards from horizontal) (see Figure 6).

**Figure 6:**
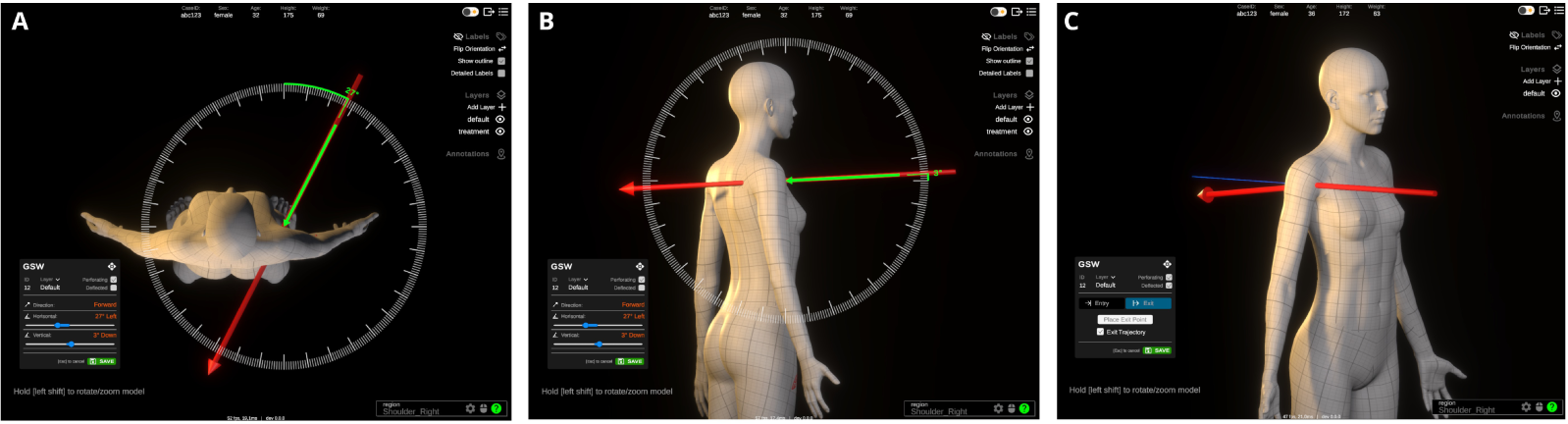
GSW trajectory placement. A) Setting horizontal trajectory angle, B) setting vertical angle, C) example deflected exit trajectory.

#### 3.4.4 Point-of-interest

Points-of-interest are placed at a single point on the body and can be used when the injury is small or of limited importance to the forensic autopsy (e.g., needle marks in diabetes) (Figure 4B).

### 3.5 Annotation data

Each new annotation is given a numerical Annotation ID (any positive number, index starting at 1). Annotation IDs can be modified after creation if annotations are incorrectly labelled, or if a different sequence of IDs is more logical to the user.

An annotation can optionally be assigned “no ID”. “No ID” annotations have their label displayed as their assigned type (e.g., abrasion, burn, intravenous line). “No ID” annotations can be used for old injuries, scars or annotations related to medical treatment. Each annotation can be assigned a ‘type’ label (e.g., abrasion, contusion, burn, laceration, etc.). Further, individual annotations can have descriptive notes added. Custom layers can be created to organise annotations logically for individual patient cases (i.e., organising burns, lacerations or blunt force trauma in individual layers). Annotations can be assigned to new layers or, in cases with few injuries, left in the default layer.

Photographs, such as those from investigative reports, or the autopsy examination, can be loaded from the user’s local disk and assigned to individual annotations. Important details within photographs can be further annotated using two-dimensional arrows (Figure 7A). Additionally, if the photo contains an object of known dimensions (e.g., a ruler), it can be calibrated to the correct scale by manually selecting two points and inputting the absolute distance between them. Calibration establishes the transformation between pixel- and absolute distance and enables the user to subsequently measure and place length markers on the photo (7B).

**Figure 7:**
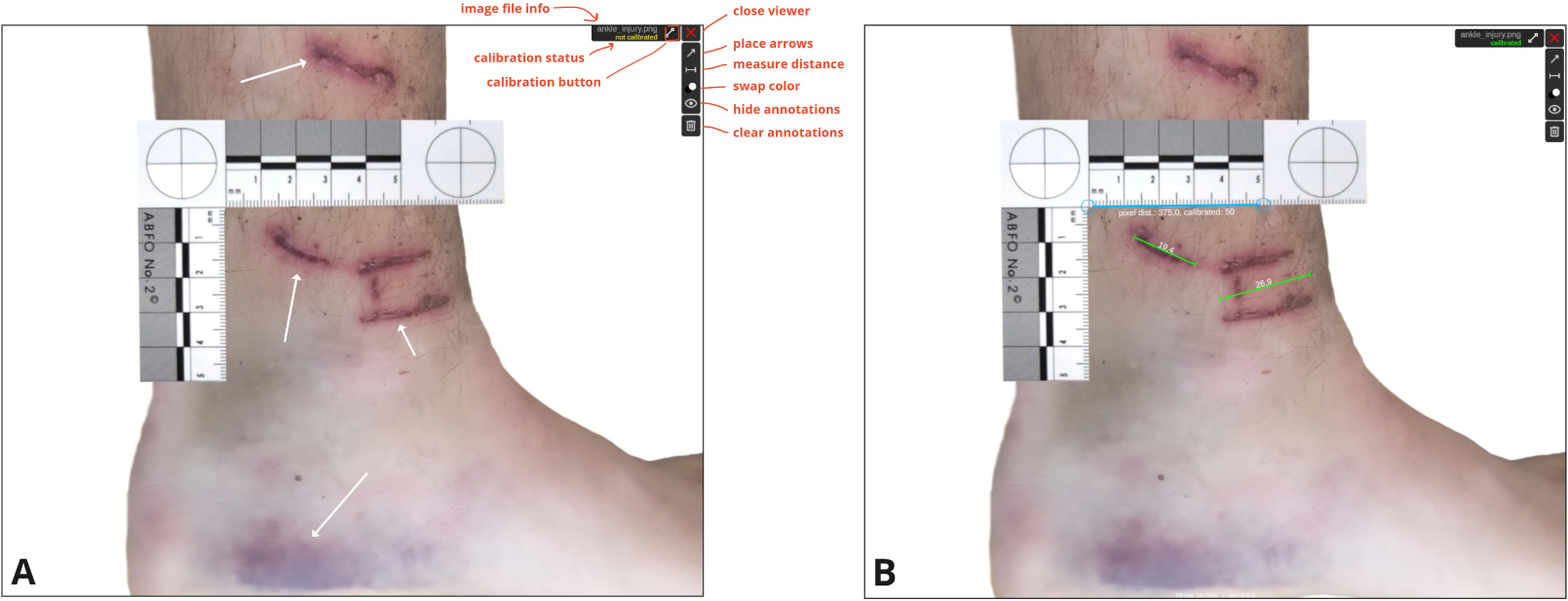
A) Example forensic photo (not actual patient data) with arrows placed to highlight important features, B) The photo is calibrated to absolute dimensions based on known object size (ruler) and measurements of absolute (in-plane) lengths are highlighted with length (mm) displayed.

When measuring length, the plane being measured (e.g., skin surface) is assumed to be on the same plane as the calibration object and the photo was taken orthogonal to the surface. Photo annotations (arrows and distance markers) can be swapped between black and white for optimal contrast given the content of the photo.

### 3.6 Save, Load and Export

#### 3.6.1 File format and saving/loading

We implemented a custom file format (.fat) allowing the user to save, load and share each case annotated in the digital tool. Previously annotated cases can be loaded for viewing or further editing by simply dragging and dropping the file onto the application window.

#### 3.6.2 Export

Cases can be exported as 2D (PDF) reports for storage alongside other case materials and investigative reports (Figure 8). The 2D export combines the front and back views into a side-by-side figure. The user can choose whether to export all annotation layers in a single figure or to create a separate figure (and page) for individual annotation layers. The figures can be exported as .png files or embedded into and exported directly as a PDF document (A4 page size). When exporting to PDF, the user can optionally import a department-specific (.pdf) template and manually mark the area of the document where the exported figures should be embedded.

**Figure 8:**
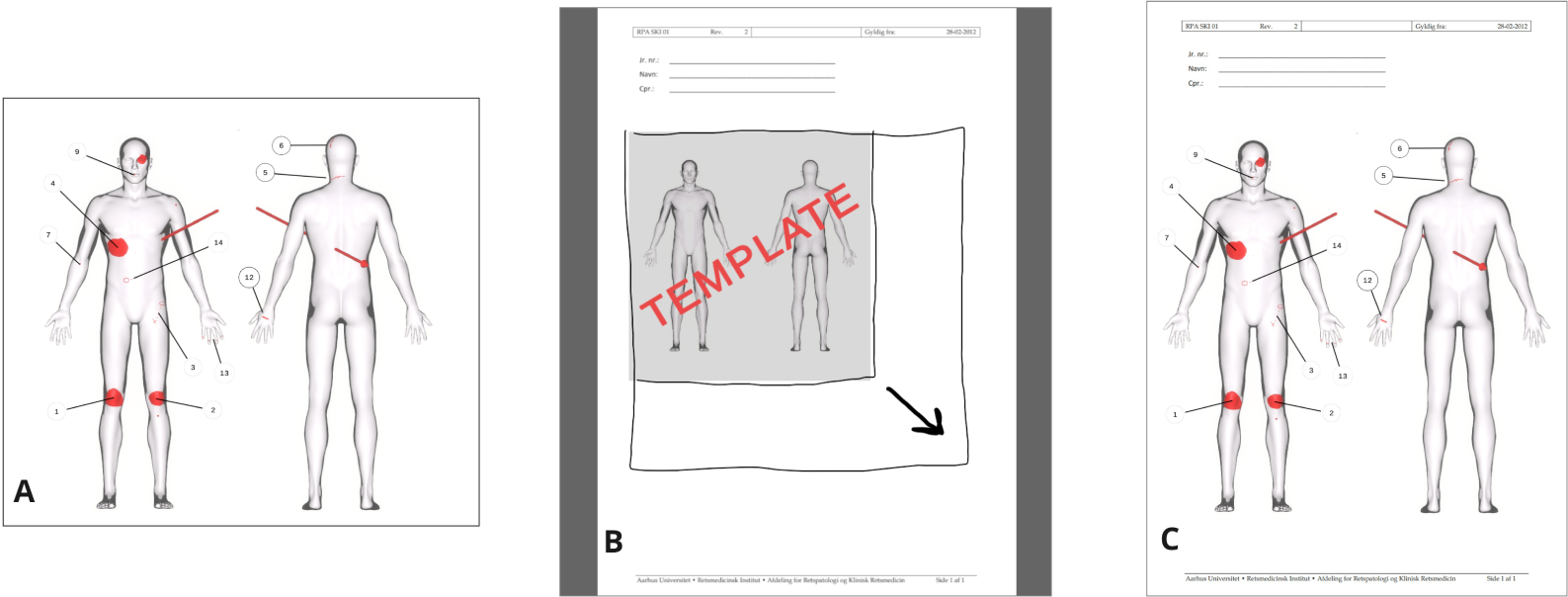
The 2D export feature allow direct export to figure or PDF. A) Front and back view with annotation labels exported as an image file (.png). B) Custom PDF template loaded in FATAL, allowing the user to manually designate size and position of the embedded FATAL figure. C) FATAL annotation figures exported directly to a PDF document.

## 4. Discussion

Here, we present the design and implementation of a digital Forensic Au-Topsy Annotation Tool (FATAL), designed to support forensic workflows. FATAL is developed iteratively with expert input from clinician and non-clinician users. Here, we describe our key findings on how traditional workflows for forensic autopsies could be supported with digital tools. Specifically, we discuss the advantages identified in moving from conventional two-dimensional paper-based formats to a three-dimensional interactive space in terms of improved accuracy, efficiency and reproducibility. Challenges for further development, from medico-legal restrictions to a small user base, are also discussed. Finally, we present possible extensions and additional use cases for digital annotation software in forensic settings.

### 4.1. Opportunities for digital tools in autopsy workflows

FATAL offers interactive 3D visualisation of standardised body models, enabling a forensic practitioner to interact with more complex reconstructions of anatomy and pathology findings than is possible using traditional paper-based 2D body diagrams. Users can zoom in on specific anatomical regions, allowing more precise annotations of both small and large injuries. Furthermore, users do not need to transform injuries from the complex three-dimensional space of the human body into a simplistic two-dimensional representation on a static figure, as used in paper-based reports. A 3D to 2D transformation becomes particularly challenging in paper-based reports, when trying to illustrate injuries that do not correspond (e.g., side or oblique viewpoints) to the static front- or back-facing figure outlines. Closer correspondence between the report space and the patient’s body should support accurate annotations [14].

Our digital tool also scaffolds annotations, supporting template-driven documentation, which has been shown to improve the consistency of reporting and replicability of findings in other domains of medicine (e.g. pharmaceutics [15]). The scaffolding provided by a digital tool has clear advantages when considering trainee forensic practitioners, and indeed, users with different training backgrounds (e.g., biology, nursing, medicine).

Further advantages of FATAL relate to the streamlining of the autopsy workflow, and the collation of relevant material into one digital tool. We developed the tool so that all forensic notes, images and other documentation can be collated into a single interface, making it easy to revisit cases, share and discuss with internal and external stakeholders.

### 4.2. Methodological lessons

Autopsy workflows have been relatively stable over the past decades, and centre on 2D paper-based reports. Interactive and immersive data visualisation prototypes in forensic contexts are only beginning to emerge (e.g., [5]), and have been far less explored than in other medical specialities such as surgery [16] and dental medicine [17]. The present iterative design process required intensive interactions between the forensic scientists and the developer, as aspects of the workflow challenges and the technical possibilities became clear to both parties. As an example of a substantial iteration, the first prototype allowed the user to draw only a circular or ellipse-shaped surface region, as shown in the example of a paper-based report. Feedback from the forensic medicine team made it clear that a circular format was overly restrictive, even if it was a typical format for paper-based annotations. The next iteration therefore implemented a feature whereby the user can draw an arbitrarily shaped (closed-loop) region.

Feedback on the next iteration clarified that the marking of a closed-loop region was insufficient for injuries such as a knife slash, longitudinal scratch marks or ligature marks. Additional functionality was implemented to allow a more diverse range of injuries to be marked (Figure 4).

### 4.3. Future directions for development

We suggest that our digital tool could be of use in communicating autopsy findings to police and other external stakeholders. While the introduction of new technologies into legal case materials is not trivial, there have been innovations towards new visualisation modalities. For example, 3D printing of skull models has been tested in the UK [18], while others have used VR to present 3D crime scenes to the authorities [19]. The challenges of introducing technology include differences in legal procedures across countries, and the necessary consideration of the emotional impact of unfamiliar visualisation modalities on jurors [20]. Nonetheless, our digital tool is well-aligned with general developments in 3D visualisations and also has the advantage of being compatible with conventional report formats, via our export PDF functionality. Further development will benefit from testing with possible external stakeholders (e.g., police, prosecutors, defence council), whose needs were not considered when designing the current tool.

We suggest a number of other functions that might be developed in future versions of the current, or a related tool. First, in the current FATAL version, annotations can only be placed on the surface of the body. Additional anatomical layers could be added allowing the user to register internal injuries (e.g., stab lacerating an organ or vessel). Second, the Abbreviate Injury Scale (AIS) [21], an anatomical coding system, could be implemented to further describe the severity of annotated injuries. Third, additional input supports could be implemented, allowing the user to make annotations using a touch screen or stylus pen. Pen inputs, for example, could allow a user experience more aligned with conventional paper formats. Fourth, additional body models could be added to match body dimensions when dealing with children or infants. Future versions might allow the user to load customised body models if preferred over the default male or female model implemented. Fifth, the exported PDFs could be extended to include interactive 3D models of annotations[22]. Finally, future development may extend to supporting 3D model printing, via export to basic 3D formats such as STL (see [23, 24]), or more complex formats, such as 3MF[25].

Forensic medicine is a relatively small medical specialty, constraining the demand and market for digital tools for autopsies. In Denmark, for example, medico-legal autopsies are infrequent, estimated to occur for between 2.4% and 2.8% of deaths [26]. In England and Wales, an average of approximately 2,000 initial forensic examinations are carried out per year [27]. While the number of direct users of FATAL is therefore limited, we anticipate other directions for development. For example, forensic medical examinations are common, relative to autopsies, and are performed in cases such as sexual assault, or other forms of physical violence, and many countries provide paper-based templates for completion by a forensic examiner (e.g., Faculty of Forensic and Legal Medicine of the Royal College of Physicians proforma examination templates [28]). The FATAL tool could be developed for forensic medical examinations, whereby diagrams of body regions are also a central component of reporting standards.

## 5. Conclusion

We initially set out to develop a digital visualisation tool to place trajectories of injuries, such as stab or gunshot wounds, on a 3D body model to export for 3D printing. During initial meetings, the clinical value of developing a more comprehensive tool than initially conceived became clear. Therefore, FATAL focuses on improving the autopsy workflow rather than providing functionality supplemental to current autopsy procedures. The significant contributions of our article are in describing the requirements for digital forensic annotation tools, as identified through an iterative, user-centred approach. We provide figures of our FATAL tool to illustrate concrete, practical functions for future developers to take into their own systems. To date, there has been minimal research on digitization and integration of tools for data visualization for forensic autopsies. Therefore, we hope that our work will serve as a springboard for further development and sharing of experiences across international contexts, given the relatively limited numbers of national users.

## Data Availability

No data available

## 6. CRediT authorship contribution statement

**Mikkel V Petersen:** Conceptualization, Methodology, Software, Visualization, Funding Acquisition, Writing – original draft. **Kasper Hansen:** Conceptualization, Investigation, Writing – review and editing. **Asser H Thomsen:** Conceptualization, Investigation, Validation, Writing – review and editing.

## 8. Acknowledgments

This work was supported by a Lundbeck Foundation grant to M.V. Petersen (R322-2019-2758).

